# Using translational *in vitro-in vivo* modeling to improve drug repurposing outcomes for inhaled COVID-19 therapeutics

**DOI:** 10.1101/2021.03.11.21253375

**Authors:** Madison Stoddard, Lin Yuan, Arijit Chakravarty

## Abstract

The ongoing COVID-19 pandemic has created an urgent need for antiviral treatments that can be deployed rapidly. Drug repurposing represents a promising means of achieving this objective, but repurposing efforts are often unsuccessful. A common hurdle to effective drug repurposing is a failure to achieve a sufficient therapeutic window in the new indication. A clear example is the use of ivermectin in COVID-19, where the approved dose (administered orally) fails to achieve therapeutic concentrations in the lungs. Our proposed solution to the problem of ineffective drug repurposing for COVID-19 antivirals is two-fold: to broaden the therapeutic window by reformulating therapeutics for the pulmonary route, and to select drug repurposing candidates based on their model-predicted therapeutic index for inhalation. In this article, we propose a two-stage model-driven screening and validation process for selecting inhaled antiviral drug repurposing candidates. While we have applied this approach in the specific context of COVID-19, this *in vitro-in vivo* translational methodology is also broadly applicable to repurposing drugs for diseases of the lower respiratory tract.

## Introduction

One year into the COVID-19 pandemic, significant progress has been made in reducing mortality due to the disease, but options for effective treatments are still extremely limited. A particular gap in the therapeutic lineup exists for early treatments, leaving watchful waiting as the paradigm for outpatient care for the disease at present [1], despite evidence that early therapeutic intervention for COVID-19 is effective [2].

Drug repurposing is a somewhat obvious strategy for rapidly expanding the range of therapeutic options for COVID-19, as it is a fast, cheap and proven methodology. Over the past decade, for example, approximately one third of all new Food and Drug Administration (FDA) approvals have originated from repurposing studies [3,4]. Accordingly, antiviral drug repurposing in the context of the COVID-19 pandemic has garnered significant attention and funding. A variety of candidate identification techniques have been pursued, including *in silico* conformational docking studies [5,6], brute force *in vitro* screening of vast libraries [7], and protein-protein interaction network analysis [8]. As a result, a wealth of *in vitro* candidate validation data is available, and some of these drugs have already been tested in COVID-19 patients.

Despite promising preclinical data and biological rationales, most of these drug repurposing efforts have met with limited clinical success, at best. For example, Remdesivir appears to confer a benefit in terms of reduced time-to-recovery, but the reduction in mortality is debatable at best [9]. As another example, hydroxychloroquine yielded no clinical benefit in large-scale clinical trials, either alone or in combination with azithromycin [10]. A third drug combination identified through repurposing efforts, lopinavir combined with ritonavir, has not been found to reduce mortality or fraction of patients with detectable viral RNA among hospitalized patients [11]. As a fourth example, ivermectin, which showed promising *in vitro* efficacy, has demonstrated marginal clinically efficacy when dosed orally [12]. Consistent with this disappointing performance in the clinic, translational modeling for ivermectin predicts insufficient exposure in the lungs under the clinically-approved oral regimen [13].

It is likely that this failure to achieve sufficient concentrations in the lungs represents a common mode of failure for systemically-dosed antivirals for COVID-19. SARS-CoV-2 infections are thought to be seeded in the upper respiratory tract, with viral particles making their way to the lungs as the disease progresses [14], and attendant lung damage is a common etiological and pathological factor for full-blown COVID-19. Therapeutic interventions that disrupt viral proliferation in the lungs thus hold the potential to impact the trajectory of the clinical disease in patients.

Despite the early failures for COVID-19, drug repurposing remains the best path forward for rapidly expanding treatment options for this disease. For other viral diseases, this approach has been effective-for example, repurposed nitazoxanide, an antiviral in extensive use for other respiratory pathogens, was found to significantly reduce the duration of influenza symptoms and reduce viral titer by 10-fold [15].

Local administration is an elegant solution to the problem of a narrow therapeutic window. For respiratory diseases affecting the lungs, inhaler- or nebulizer-administered therapies represent a safe and efficient means of delivering drug to the active site [16]. Many well-characterized technologies are already available for drug inhalation, and some antiviral drugs are already administered through this route [17]. Recent publications have proposed the pulmonary route as a repurposing strategy for COVID-19, focusing on drugs with prior clinical data via the pulmonary route [18] or assessing lung concentrations for therapeutics administered via the approved systemic routes [19].

In this work, we have sought to extend this framework further by asking the question: “are there drugs for which a pulmonary route represents a viable clinical strategy, with reformulation if necessary?” To answer this question, we have developed a two-stage model-based screening platform to identify promising candidates among *in vitro*-validated drugs which have undergone clinical testing for another indication. In the first stage, we employ a rapid computational screen to select *in vitro*-characterized drugs likely to exhibit a high therapeutic index (TI) in the inhaled setting, including both approved drugs with prior clinical experience via the pulmonary route and drugs for which the pulmonary route would require reformulation and redevelopment. Next, we built a computational deposition-PK model to refine the therapeutic index calculation for candidates predicted to have a wide TI and provide preliminary lung kinetics estimates. This two-step process provides *in silico* proof-of-concept for a pulmonary delivery approach and supports the rational design of dosing regimens in the new indication. Such an approach allows us to increase the likelihood of achieving therapeutic drug concentrations for COVID-19 treatment while avoiding dose-limiting toxicity.

## Methods

### Rapid drug screen based on estimated therapeutic index

We propose a two-stage repurposed drug screening and *in silico* validation process based on maximizing TI for inhalation. The purpose of the first stage is to rapidly screen a large number of repurposing candidates for likely large TI. This screening process is summarized in Figure 1.

**Figure 1.**
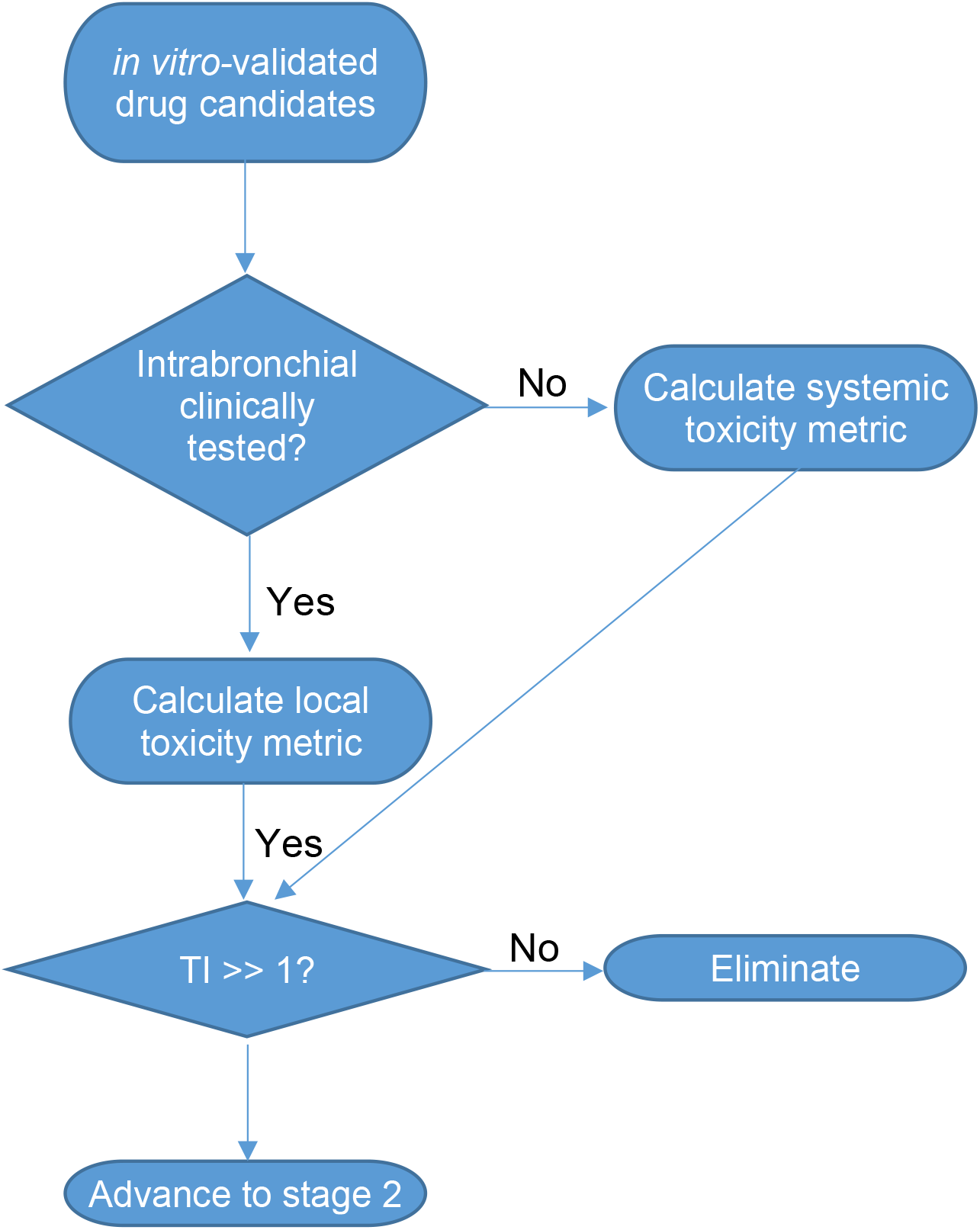
Flow-chart for rapid first-pass screen of *in vitro*-validated drug candidates for repurposing in the inhaled setting. A therapeutic index is calculated based on local or systemic toxicity and used to select drugs expected to have a wide therapeutic window after inhalation.

Repurposing candidates with clinical precedent as inhaled therapeutics is the most desirable approach as in this case the maximum tolerated dose (MTD) via the inhalation route is known. However, repurposing candidates lacking clinical precedent as inhaled therapeutics can be evaluated on the basis of systemic toxicity. We have outlined both approaches. For the purpose of this screen, all repurposing candidates must have *in vitro* antiviral activity data yielding an EC_90_ (90% maximal effective concentration). Commonly, EC_50_ (half-maximal effective concentration) values are reported in the literature. The EC_90_ is preferably estimated from the *in vitro* dose-response curve, but may also be estimated using the following equation:

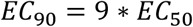

This relation assumes a Hill coefficient of 1. A Hill coefficient of 1 is typical for a small molecule that binds a target with a single binding site without cooperativity [20]. However, some antivirals may exhibit distinct binding and potency properties, so estimating the EC_90_ based on *in vitro* data will yield a more accurate result.

Based on the EC_90_, a simple therapeutic index (TI) calculation can be performed based on the highest known tolerated dose. If the MTD for the inhaled settfing is available, the following local TI calculation can be made:

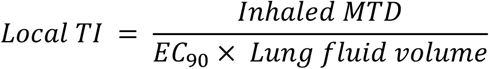

This preliminary TI calculation represents the ratio of the expected lung concentration at the inhaled MTD to the experimental EC_90._ The fluid volume of the lungs is approximately 25 mL [21]. For the purpose of this calculation, we assumed that all administered drug reaches the lung (perfect deposition). This is optimistic but suitable for a preliminary screen. The extent of deposition is refined in the candidate validation stage.

If clinical toxicity data for inhalation is not available for a candidate drug, a systemic TI can be estimated based on the systemic MTD and peak plasma concentration (C_max_). This calculation is as follows:

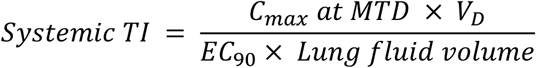

where V_D_ represents the plasma volume of distribution. We note that for a drug administered by IV bolus, the C_max_ at MTD multiplied by V_D_ is equivalent to the MTD. This calculation ensures that the estimated inhaled therapeutic dose would not cause intolerable systemic toxicity assuming that the drug transits instantaneously from the lungs to systemic circulation. This is a pessimistic assumption but covers the potential for highly rapid absorption from the lungs [22].

### Candidate validation and go/no-go based on lung deposition-pharmacokinetic model

The second stage of the drug repurposing platform refines the therapeutic index calculation and provides a preliminary lung pharmacokinetic model. In this stage, we estimated the fractional lung deposition of drug particles administered via the pulmonary route, for a number of different candidate inhalation devices (nebulizers and inhalers), converted this fractional deposition into a bolus or rate of drug entry into the lungs, and built a simple compartmental PK model to predict the concentration of drug in the lungs over time. This allows us to estimate the therapeutic dose required to achieve the EC_90_ in the lungs. The ratio of the inhaled maximum tolerated dose to the predicted therapeutic dose is the model-based refined local TI. We used this more time-intensive method to verify promising drug repurposing candidates with a wide TI. This process is outlined in Figure 2.

**Figure 2.**
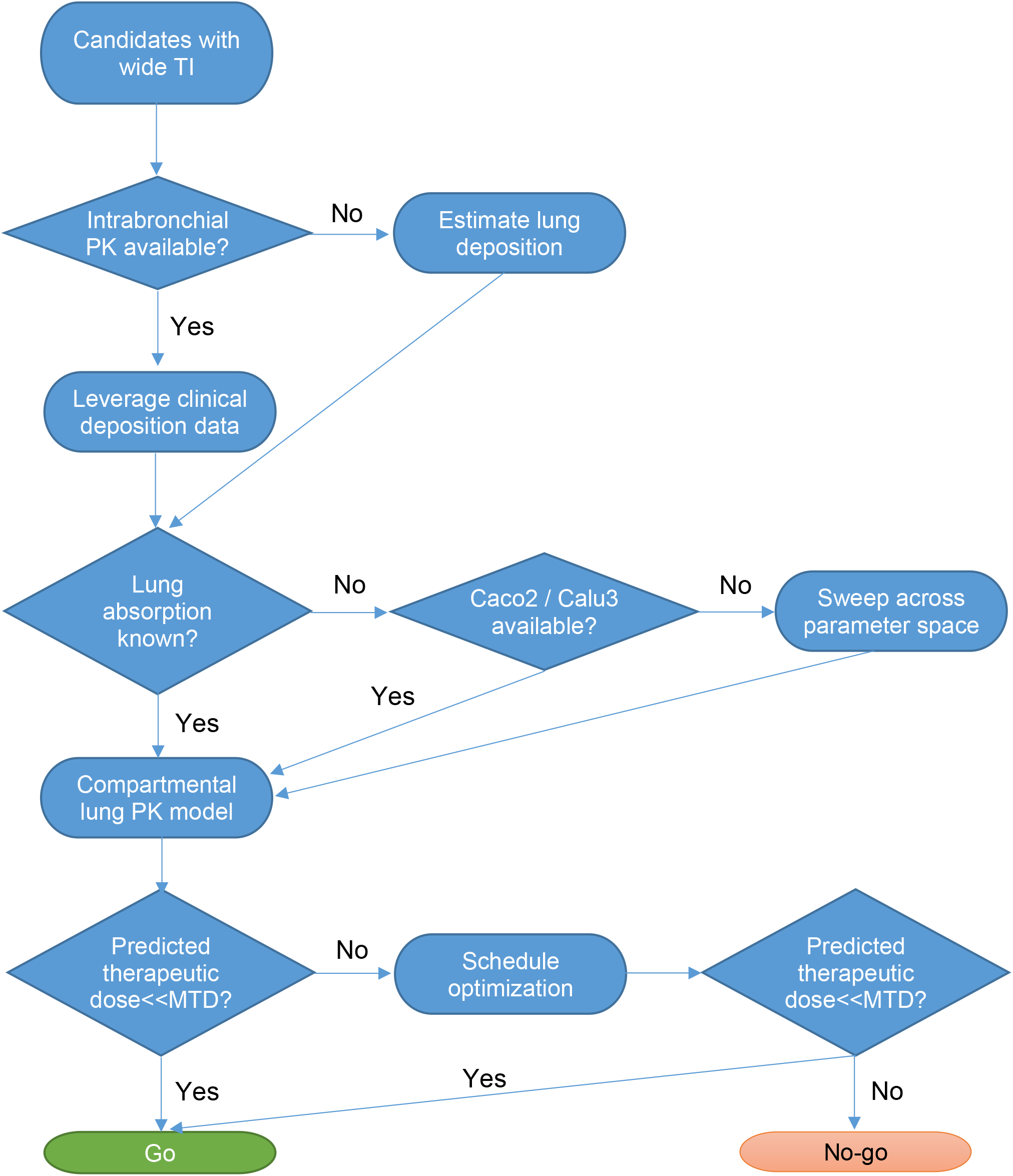
Flowchart outlining decision process based on a refined TI calculation. Lung deposition and absorption are estimated for the drug of interest to allow model-based prediction of the therapeutic dose. Drugs suitable for repurposing will have a therapeutic dose that is significantly less than the maximum tolerated dose.

For some candidate drugs, lung deposition data based on prior clinical studies may be available. Where possible, we have leveraged this data. In the absence of clinical data, we estimate lung deposition based on the known particle distribution of the intended inhalation device and the known fraction of particles reaching the lungs based on their size. Computational fluid dynamics simulation of lung deposition represents a high-fidelity alternative which is outside the scope of this paper [23].

In this text, we have used ribavirin as an example candidate for repurposing in COVID-19. Under its current FDA approval, ribavirin is administered by Small Particle Aerosol Generator (SPAG-2) at a concentration of 20 mg/mL [24]. The SPAG-2’s particle size distribution is well-characterized [25]. We used the known relationship between particle size and fractional deposition in the alveolae and bronchi to estimate the fraction by mass of each particle size bin in the SPAG-2 emission distribution that reaches the lungs [26]. The data for the SPAG-2 nebulizer is summarized in Table 1. We summed over the particle size distribution bins to determine the total fractional mass reaching the lungs. Autopsy studies have demonstrated direct viral cytopathic effect in both the alveolae and the bronchi, indicating the relevance of both sites [27].

**Table 1.** Data for estimating fractional lung deposition of SPAG-2 nebulizer.

| Particle diameter (um) | % of emission by mass | % bronchial deposition | % alveolar deposition |
| --- | --- | --- | --- |
| >9 | 2 | 15 | 1 |
| 5.8 - 9 | 0.6 | 29 | 4 |
| 4.7 - 5.8 | 0.9 | 34 | 23 |
| 3.3 - 4.7 | 4.8 | 30 | 32 |
| 2.1 - 3.3 | 16.4 | 19 | 39 |
| 1.1 - 2.1 | 22.4 | 8 | 35 |
| 0.7 - 1.1 | 20.4 | 3 | 23 |
| 0.4 - 0.7 | 18.2 | 2 | 13 |
| <0.4 | 14.2 | 2 | 11 |

The lung deposition model serves as the drug input to the lung compartmental PK model. For a nebulized formulation, we determined the rate of drug entry into the lungs and treated this as a zero-order input over the administration duration. For formulations delivered by inhaler, the bolus dose reaching the lungs is treated as the initial condition and is calculated as the total dose multiplied by the fraction of drug deposited in the lungs. For nebulized drugs, we calculated the rate of drug entry as follows:

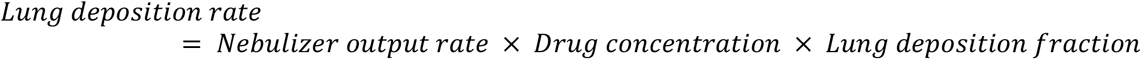

For ribavirin, the output rate of the SPAG-2 nebulizer is 12 mL/hr [25], and in the FDA-approved regimen, a concentration of 20 mg/mL is administered [24].

The lung PK model is comprised of a single compartment representing the lungs. We chose a one-compartment model for the general case because for most drugs, insufficient data is available to support a more complex model. The model accounts for the input of drug from the nebulizer or inhaler and the first-order loss of drug from the lungs through metabolic breakdown or absorption to systemic circulation. For most drugs, metabolic breakdown in the lungs is minimal relative to the rate of absorption to systemic circulation [28]. We assume that the deposited drug distributes instantaneously into the lung fluid volume of 25 mL [21]. The one-compartment model is described by the following ordinary differential equation (ODE):

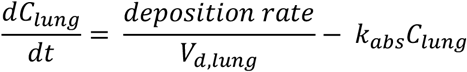

where C_lung_ is the concentration of drug in the lung, V_d, lung_ is the lung fluid volume, and k_abs_ is the rate of drug clearance from the lung. For drugs administered by inhaler, the drug reaches the lungs as a bolus. In this case, the deposition rate is set to zero and the following initial condition is set:

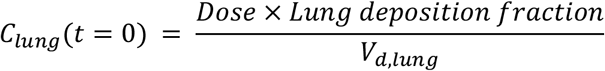

For some drugs, the half-life in the lungs or the rate of absorption to systemic circulation is known from human pharmacokinetic studies. To predict the lung PK of ribavirin, we have relied on literature estimates for the rate of drug clearance from the lungs. Based on prior PK studies, the half-life of ribavirin in the lungs is 2 hours [29]. In the absence of such data, we estimated the rate of absorption based on Caco2 or Calu3 permeability reported in the literature [30]. The absorption rate can be estimated by multiplying the flux across the Caco2 cell monolayer by the surface area of the lungs [31]. The surface area of the adult human lungs is estimated to be 70 m^2^ [21].

This combined deposition-lung PK modeling framework allows translation of *in vitro* effective concentrations to *in vivo* predicted therapeutic doses. Determining effective dose requires knowledge of the PK driver of efficacy for antivirals. Commonly, peak concentrations at the active site (C_max_) or trough concentrations between doses (C_min_) have been found to drive efficacy of antivirals [32,33]. Conservatively, we have set C_min_ ≥ EC_90_ as the constraint for the effective dose. This reduces the likelihood that subtherapeutic exposure will permit evolved resistance in the lungs [34]. Depending on practical constraints, once-daily or multiple daily administrations may be feasible. Thus, the dosing schedule and dose may both be optimized to achieve effective concentrations. Once the total daily effective dose for a given schedule has been determined, the refined local TI can be calculated as the ratio of the daily MTD to the effective dose.

## Results

### Many systemically administered repurposed drugs are likely to face narrow TIs for COVID-19

Consistent with work done by others [35], we note that therapeutically active systemic concentrations are not achieved for many COVID-19 repurposing candidates after oral or intravenous administration (Table 2). This is indicated by a systemic administration TI less than 1 (for example, ribavirin and azithromycin). For many other candidate drugs, the TI is near 1, indicating little room for translation error or higher dosing to achieve therapeutic concentrations for a longer duration (for example, teicoplanin and indomethacin). The ability to achieve effective concentrations at the MTD is a significant constraint for systemically administered COVID-19 repurposing candidates.

**Table 2:** Feasibility of achieving EC_90_ by systemic administration.

| Drug | Plasma C<sub>max</sub> at MTD (μM) | EC90 (μM) | Systemic administration TI |
| --- | --- | --- | --- |
| Aprotinin | 250 KIU / mL* [36] | 250 KIU/mL [37] | 1 |
| Camostat | 0.2 [38] | 5 [39] | 0.034 |
| Indomethacin | 5.6 [40] | 3.8 [41] | 1.47 |
| Teicoplanin | 58.5 [42] | 50 [43] | 1.17 |
| Niclosamide | 2 [44] | 1.2 [45] | 1.67 |
| Azithromycin | 0.7 [46] | 8.7 [47] | 0.077 |
| Ribavirin | 11.3 [48] | 304 [9] | 0.037 |
| Dipyridamole | 7 [49] | 4.8 [50] | 1.46 |

\*Aprotinin is measured in kallikrein inhibitor units (KIU).
Ciclesonide is excluded from this analysis because administration is almost exclusively by inhaler.

### Systemic toxicity is expected to be significantly reduced by local inhaled administration

Although a wide range of TIs are predicted by our simple metric for systemic tolerability after inhalation, all TIs predicted are larger than 10 (Table 3). This indicates that systemic toxicity is unlikely to be a constraint for any of these drug candidates if administered by inhalation in COVID-19 patients. Notably, the therapeutic indices for inhalation for each drug are much wider than the corresponding indices for systemic administration (Table 2). This emphasizes the importance of local administration in drug repurposing for COVID-19.

**Table 3:** Systemic toxicity TI for COVID-19 repurposing candidates.

| Drug | MTD plasma C<sub>max</sub> (μM) | Plasma V<sub>D</sub> (L) | EC90 (μM) | Systemic toxicity TI |
| --- | --- | --- | --- | --- |
| Aprotinin | 250 KIU / mL* [36] | 26.5 [51] | 250 KIU/mL [37] | 1,060 |
| Camostat | 0.2 [38] | 30.4 [38] | 5 [39] | 41 |
| Indomethacin | 5.6 [40] | 27.2 [52] | 3.8 [41] | 1,600 |
| Teicoplanin | 58.5 [42] | 4.6 [53] | 50 [43] | 262 |
| Azithromycin | 0.7 [46] | 1,840 [54] | 1.2 [47] | 5,650 |
| Ribavirin | 11.3 [48] | 2,000 [55] | 8.7 [9] | 2,960 |
| Dipyridamole | 7 [49] | 92 [56] | 4.8 [50] | 5,370 |
| Ciclesonide | 0.0025 [57] | 9,800 [58] | 6.3 [59] | 157 |

\*Aprotinin is measured in kallikrein inhibitor units (KIU).
Limited PK data is available for niclosamide, so the systemic toxicity TI calculation was not performed.

### For most drugs studied, tolerability after local administration allows a wide TI

In Table 4, we calculated therapeutic indices based on local tolerability (MTD for inhaler- or nebulizer-administered formulations) for 9 drugs for which a dose-tolerability data was available in the context of inhalation. All drugs studied appear suitable for inhaled or nebulized administration in COVID-19 with the exception of aprotinin, which is not expected to reach therapeutic concentrations in the lungs at the highest tolerated inhaled dose.

**Table 4:** Local toxicity TI.

| Drug | Inhaled MTD (mg per day) | EC90 (μM) | Local toxicity TI |
| --- | --- | --- | --- |
| Aprotinin | 160 KIU* [60] | 250 KIU/mL [37] | 0.026 |
| Camostat | 1.6 [61] | 5 [39] | 26 |
| Indomethacin | 50 [62] | 3.8 [41] | 1,470 |
| Teicoplanin | 200 [63] | 50 [43] | 85 |
| Niclosamide | 50.5 [64] | 1.2 [45] | 5,130 |
| Azithromycin | 70 [65] | 8.7 [47] | 430 |
| Ribavirin | 6,000 [66] | 304 [9] | 3,240 |
| Dipyridamole | 50 [67] | 4.8 [50] | 825 |
| Ciclesonide | 2.88 [68] | 6.3 [59] | 34 |

\*Aprotinin is measured in kallikrein inhibitor units (KIU).

We also note that for most drugs studied, the MTD for local administration is a more significant constraint than the highest tolerated plasma C_max_ (for either local or systemic delivery, whichever is higher). Thus, the availability of clinical data for inhalation is highly desirable for estimating the therapeutic window. This also justifies the choice to limit PK prediction in the validation step to the lungs rather than attempting to predict plasma concentrations after inhalation.

### Refined local TI calculation based on deposition-PK model

To demonstrate the validation process, we calculated a more precise TI for ribavirin, a clinically approved antiviral drug proposed for repurposing in COVID-19. First, we estimated the fractional deposition in the lung based on the data summarized in Table 1. Based on this data, we estimated total lung deposition to be 33.1% of the total nebulized dose, with 8.4% of the dose deposited in the bronchial space and 24.7% deposited in the alveolae.

Our objective was to maintain ribavirin concentrations in the lungs above the EC_90_ at all times (C_min_ ≥EC_90_). Total dose and dosing schedule are both determinants of C_min_, so we tested three possible administration schedules: daily (Figure 3A), twice-daily (BID, Figure 3B), and three times daily (TID, Figure 3C). These figures show the concentration-time profile in the lungs for a ribavirin regimen where the nebulized concentration is 20 mg/mL (consistent with the approved regimen) and the duration of nebulization is set to the shortest duration for which C_min_ ≥EC_90._

**Figure 3.**
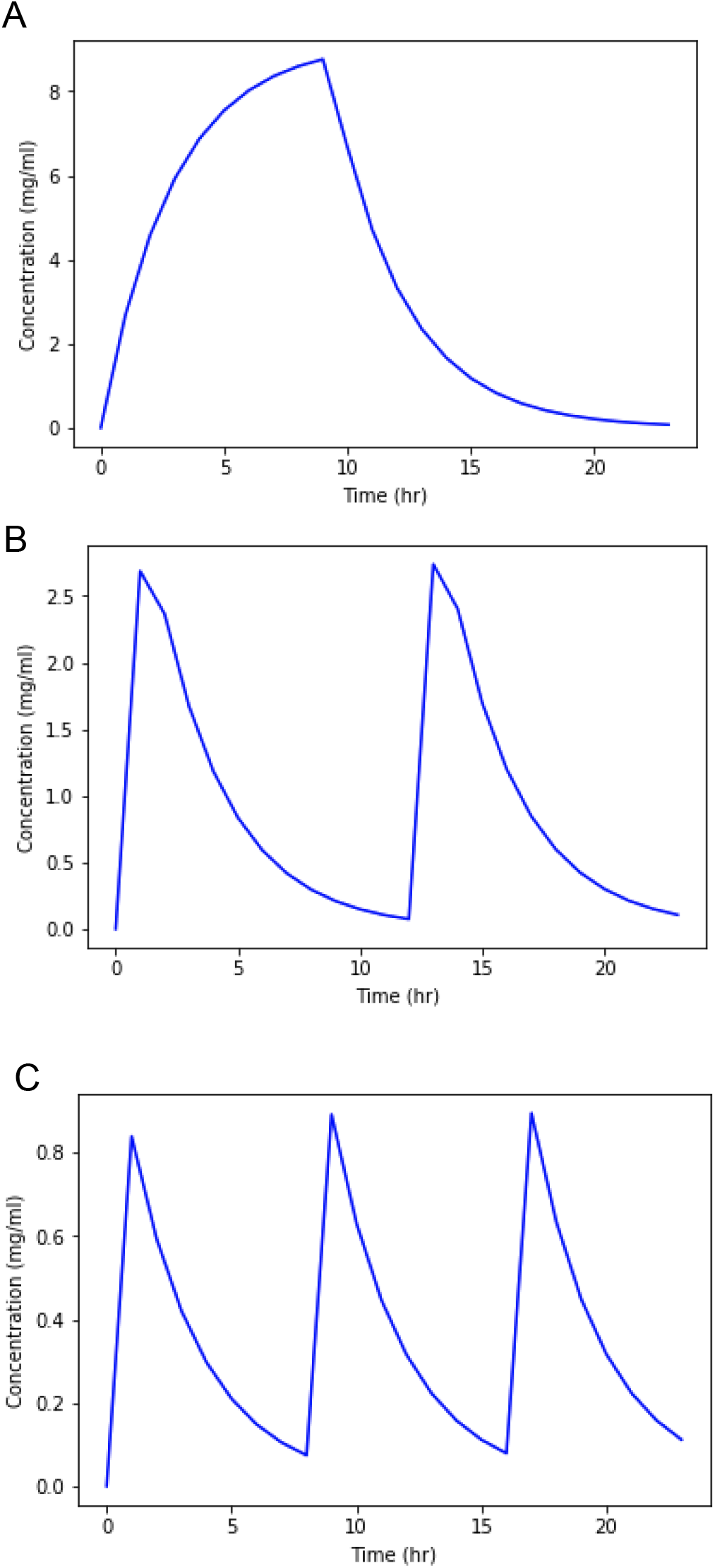
Model-predicted lung pharmacokinetics for nebulized ribavirin. Predicted lung concentration-time profiles for A) once-daily, B) twice-daily (BID), or C) three times daily (TID) administration by SPAG-2 nebulizer. Nebulization duration is optimized such that the minimum lung concentration (C_min_) is greater than or equal to the EC_90_ for ribavirin (0.074 mg/mL).

Table 5 summarizes the parameters and TIs of these three possible regimens.

**Table 5.** Comparison of possible ribavirin administration regimens.

| Number of sessions daily | Duration of each session (hrs) | Total dose administered (mg) | TI |
| --- | --- | --- | --- |
| 1 | 9.2 | 2,208 | 2.72 |
| 2 | 1.2 | 576 | 10.4 |
| 3 | 0.35 | 253 | 23.7 |

Thus, multiple administrations can raise the C_min_. We note that these therapeutic indices are much smaller than the simple local TI metric outlined in Table 4. The metric in Table 4 reflects the therapeutic index defined with respect to achieving a lung C_max_ greater than or equal to the EC_90_; clearly, more drug is required and kinetic considerations must be made to maintain concentrations greater than EC_90_ at all times.

## Discussion

Fully one year into the COVID-19 pandemic, a dearth of effective antiviral treatments persists [69]. Drug repurposing is a promising avenue for rapidly advancing well-characterized and affordable treatments to market and has already been pursued extensively in COVID-19. Consequently, the *in vitro* activity of many drugs against SARS-CoV-2 has been demonstrated in the literature [70]. Many of these candidates have advanced to clinical trials in COVID-19 [71]. However, so far this has translated to limited success in the clinic.

In this work, we hypothesized that a major obstacle to successful treatment of COVID-19 is a lack of a therapeutic window. In Table 2, we show that many antiviral COVID-19 repurposing candidates do not achieve therapeutic concentrations at the target site upon systemic administration at the MTD. To address this challenge, we have described and implemented an efficient screening and validation process for repurposing clinically tested drugs for the pulmonary route of administration. Our rapid computational screen is designed to allow candidate prioritization based on publicly available *in vitro* efficacy and clinical tolerability data. This method focuses on maximizing the therapeutic index for COVID-19 via the pulmonary route, which we and others have identified as a key determinant of success for repurposed drugs [13].

An elegant solution to the problem of a narrow therapeutic window after systemic administration is to deliver drug directly to the active site, which in the case of COVID-19 therapeutics is the lung. Our work here shows that for this set of clinically characterized repurposing candidates, systemic toxicity will be dramatically reduced for nebulized or inhaled formulations compared to oral or intravenous administration, allowing therapeutic concentrations to be achieved in the lungs. For most of these drugs, we also demonstrate that the known inhaled or nebulized tolerated dose is likely to produce a C_max_ in the lungs in excess of the *in vitro* EC_90_.

Finally, we demonstrate a lightweight *in vitro*-*in vivo* translational modeling framework to validate drug repurposing candidates for therapeutic inhalation in COVID-19. This approach allows drug selection with confidence that a therapeutic window is available and supports further dose and schedule optimization. Our work confirms that nebulized ribavirin is a promising candidate for COVID-19 therapy and demonstrates that a BID or TID schedule is suitable for achieving sustained therapeutic concentrations without inducing local or systemic toxicity.

## Data Availability

The work describes a mathematical methodology based on publicly available data. All data used in the work was sourced from prior publications.

## Notes

### Competing Interest Statement

MS, LY and AC are shareholders and employees of Fractal Therapeutics Inc. This work was funded by Fractal Therapeutics. Fractal Therapeutics does not benefit in any way from sharing the work here (strategy for identifying new antiviral combinations) with potential competitors. 

### Funding Statement

LY and MS's salaries were paid by Fractal Therapeutics.

